# The support needs of Australian primary health care nurses during the COVID-19 pandemic

**DOI:** 10.1101/2020.06.19.20135996

**Authors:** Elizabeth Halcomb, Anna Williams, Christine Ashley, Susan McInnes, Catherine Stephen, Kaara Calma, Sharon James

## Abstract

**Aim:** To identify Australian primary healthcare nurses immediate support needs during the COVID-19 pandemic.

**Background:** COVID-19 has had widespread implications for primary healthcare nurses. Supporting these nurses’ capacity to deliver quality care ensures that ongoing health needs can be met.

**Methods:** Primary healthcare nurses were recruited to an online survey via social media and professional organisations in April 2020.

**Results:** Six-hundred and thirty-seven responses were included in analysis. Participants provided 1213 statements about perceived supports required to provide quality clinical care. From these, seven key categories emerged, namely; personal protective equipment, communication, funding, industrial issues, self-care, workplace factors and valuing nurses.

**Conclusion:** A number of key issues relating to personal health and safety, care quality, and job security need to be addressed to support primary healthcare nurses during the COVID-19 pandemic. Addressing these support issues can assist in retaining nurses and optimising the role of primary healthcare nurses during a pandemic.

**Implications for nursing management:** Responding to the needs of primary healthcare nurses has the potential to facilitate their role in providing community based healthcare. This knowledge can guide the provision of support for primary healthcare nurses during the current pandemic, as well as informing planning for future health crises.

## 1 INTRODUCTION

The world has experienced several infectious respiratory viruses in recent years, including severe Acute Respiratory Syndrome (Maunder, 2004), novel influenza A / H1N1 (swine flu)(Fitzgerald, 2009) and Middle East Respiratory Syndrome (Kim, 2018). However, nothing has compared to the scale with which the novel Corona Virus Disease 2019 (COVID-19) pandemic has swept the globe (Huang & Rong Liu, 2020). A pandemic is a global outbreak of a disease, that creates a public health crisis and commonly results in a high rate of mortality and socioeconomic disruption (Young & Fick, 2020).

Any pandemic has a significant impact on health systems, particularly the workforce (Ives et al., 2009; Seale, Leask, Po, & MacIntyre, 2009). Nurses constitute the largest professional group within healthcare globally and, as such, are a pivotal component of coordinated responses to such events (Jordan, Adab, & Cheng, 2020). Historically, nurses have always played a role in infection prevention and control, and in caring for people during crises (Corless et al., 2018). The COVID-19 pandemic has been no exception. Nurses have played a key role, working both to identify, isolate and manage those with COVID-19 and supporting those who have non-COVID-19 related health needs (Halcomb et al., 2020).

Of necessity, when pandemics occur, the predominant focus of health systems is to build the capacity of acute hospital services to care for the critically ill (Young & Fick, 2020).

However, much health care is delivered outside of hospital settings by primary health care (PHC) nurses. PHC nurses work in a range of diverse settings including general (family) practice, community nursing services, schools, residential care settings and within workplaces (Guzys, Whitehead, Brown, & Halcomb, 2017). During pandemics PHC nurses play a vital role in supporting people in the community to maintain health through education around infection control and promoting access to ongoing health services in the community, including preventative activities (e.g. vaccinations) and chronic disease management (McMillan, 2020). Without this continuation of care, there is likely to be a secondary mortality and morbidity related to health service disruption and delayed access to care.

It has been acknowledged that the PHC workforce could be quickly overwhelmed in the event of a pandemic (Shaw, Chilcott, Hansen, & Winzenberg, 2006). Despite the clear role of PHC nurses in responding to a pandemic, there is limited evidence regarding the nurses preparedness to respond to a pandemic or the supports required to facilitate high quality clinical nursing care (Shaw et al., 2006). The aim of the overall study was to explore the experiences of Australian PHC nurses during COVID-19 in a timely way to inform emerging policy, nursing management and clinical practice. This paper seeks to report the immediate support requirements identified by Australian PHC nurses to enable them to provide quality care during the COVID-19 pandemic.

## 2 METHODS

This study used an online cross-sectional survey delivered by Survey Monkey (SurveyMonkey Inc., nd) to collect data.

### 2.1 Instrument

A survey tool was purposefully designed by the research team based on the literature and observations related to the unfolding pandemic impact on PHC nursing. The survey tool consisted of quantitative items concerned with participant demographics, employment conditions, knowledge, attitudes and supports, current service provision, COVID-19 testing, personal protective equipment. In addition, text boxes were provided to allow participants to provide in-depth qualitative responses. Prior to survey distribution, the survey was reviewed by six individuals including nurse academics, policy experts and PHC nursing clinicians. Minor modifications to some aspects of the wording and format of the tool occurred as a result of feedback.

This paper reports the findings associated with one of the in-depth qualitative open-ended responses that asked participants to *“suggest the top three things that can be done to support you to continue to provide quality care in the community during the COVID-19 pandemic”*. The quantitative findings of the survey have been reported separately due to the depth of data (Halcomb et al., 2020).

### 2.2 Participants

Baccalaureate prepared (or equivalent) registered nurses, diploma prepared enrolled nurses or Masters prepared Nurse Practitioners that were currently working in an Australian primary health care setting were eligible to participate in the study.

### 2.3 Data collection

The survey was open between April 9 to April 20, 2020. This short timeframe was used to capture a snapshot of the situation and facilitate rapid reporting to inform policy making and clinical practice. Recruitment relied on convenience and snowballing methods via distribution of the survey link through social media (LinkedIn, Facebook, Twitter) and key organisations (Australian Primary Health Care Nurses Association, Australian College of Nursing and Primary Health Networks) who were asked to disseminate study information via their email and social media channels. The survey link took potential participants to an information sheet outlining the purpose of the study and use of data before they entered the online survey tool.

### 2.4 Ethical considerations

The Human Research Ethics Committee at the University of Wollongong (Approval Number HE2020/161) approved the study. This approval was endorsed by the University of Notre Dame, Sydney (Approval Number 2020-056S). Participants were guaranteed confidentiality as survey responses were not identifiable.

### 2.5 Data analysis

The survey data were downloaded from SurveyMonkey Inc. (nd) into SPSS Version 23 (IBM Corp., Released 2015) for analysis. The dataset were then checked for missing data (>50% missing data) or where the participant did not meet the inclusion criteria (either due to not being a registered / enrolled nurse or not working in PHC), which were then excluded from analysis.

Qualitative data were exported to Microsoft Excel (Microsoft Corporation, 2016) and analysed using a broad thematic analysis approach based on a number of categorising strategies. Responses were compared and initially coded into a broad list of main organisational categories that defined the key areas in which supports were identified as being required. The main organisational categories were further cross compared and a number of elements related to each main organisational category were established. These elements provided a detailed description of the type and extent of supports perceived by the participants to be necessary in order to provide quality care during the COVID-19 pandemic. This process was initially undertaken by one researcher (XX) and cross-checked by a second researcher (XX). Discrepancies in the labelling of the key organisational categories and subsequent labelling and allocation of the elements to an organisation category were discussed and resolved by the team.

## 3 RESULTS

### 3.1 Demographics

Ninety eight of the 735 responses received were excluded as they had either missing data or did not meet the inclusion criteria, leaving 637 responses included within the analysis. Just over half of participants worked in general practice (n=351; 55.1%), with 106 (16.6%) participants employed as community nurses and 180 (28.3%) employed in various other PHC roles, including schools, universities and Aboriginal medical services. Most participants were female (n=613; 96.2%) and employed as Registered Nurses (n=555; 87.1%). The participants were highly experienced, with over half (n=338; 53.1%) having been employed in nursing for over 21 years. The mean duration of experience working in PHC was 10.7 years.

### 3.3 Support Needs

Four hundred and sixty (72.4%) participants provided a total of 1213 statements regarding supports required to provide quality care during the COVID-19 pandemic. Coding these statements revealed seven major categories based on counts of the number of statements in each category (Table 1). These categories were; personal protective equipment, communication, funding, industrial issues, self-care, workplace factors and valuing nurses. With the exception of the category related to personal protective equipment, where participants simply wanted better access to this equipment, each support category was found to have a number of elements.

**Table 1.** Key Categories

| Category | n (%) | Category | n (%) |
| --- | --- | --- | --- |
| **Personal Protective Equipment** |  | **Self-care** |  |
| **Total** | **355 (29.3%)** | **Total** | **49 (4%)** |
| **Communication** |  | Mental health | 28 |
| **Total** | **255 (21.0%)** | Staying well | 7 |
| Protocolised care | 62 | Time | 6 |
| Delivery of information | 57 | Personal safety | 6 |
| Professional Education | 52 | Abuse | 2 |
| Community Messaging | 44 | **Workplace Support** |  |
| General Information | 24 | **Total** | **45 (3.7%)** |
| Information on Services | 8 | Relations with Management | 19 |
| Other | 8 | Workplace Communication | 12 |
| **Funding of Services** |  | Work practices | 5 |
| **Total** | **134 (11.0%)** | Other | 5 |
| Nurse telehealth item | 62 | Education | 2 |
| Nurse billing | 36 | Teamwork | 2 |
| Fund services | 13 | **Valuing Nurses** |  |
| Job security | 8 | **Total** | **38 (3.1%)** |
| Nurse Practitioner | 3 | Acknowledgement | 16 |
| **Industrial Issues** |  | Need to be included | 5 |
| **Total** | **123 (10.1%)** | Recognition of risk | 5 |
| Job security | 27 | Understanding the Role | 4 |
| Additional staffing | 16 | Treatment of Nurses | 4 |
| Fair pay | 13 | Nursing leadership | 4 |
| Leave / Leave Cover | 16 | **Other Minor Categories** |  |
| Rewards / Incentives | 18 | **Total** | **214 (17.6%)** |
| Other | 10 | Staffing | 35 |
| Funding of Jobs | 8 | Testing | 34 |
| Jobseeker | 8 | General support | 25 |
| No current employment | 7 | Telehealth | 24 |
|  |  | Influenza vaccine | 19 |
|  |  | Equipment | 14 |
|  |  | Pandemic measures | 14 |
|  |  | Working from home | 12 |
|  |  | Cleaning | 9 |
|  |  | Other | 28 |

#### 3.2.1 Personal Protective Equipment

Over three quarters of statements (77.2%) identified the need for an adequate supply of personal protective equipment to enable the provision of quality routine care during the pandemic. *“Access to face shields, masks, Gowns is a high priority”*. Participants discussed the impact of worldwide shortages of personal protective equipment on workplace supply. Additionally, participants highlighted a need to have clear protocols about the appropriate use of personal protective equipment communicated in order to guide practice and promote consistency of use between health professionals. Participants identified the need for *“adequate PPE* [*personal protective equipment*] *supplied by the government not at a cost to the Practice”* for small businesses such as general practices, particularly in an environment where costs have become *“expensive, inflated because of demand”*.

#### 3.2.2 Communication

Just over half of the statements (55.4%) referred to the need for high level communication supports in order to continue to provide quality nursing care. Communication elements where supports were required included standardised protocols to govern clinical care, regular and up to date delivery of information related to COVID-19, and access to COVID-19 education for both health professionals and the general community.

In terms of communication delivery about the pandemic, it was seen as important for “*continuous up to date information*” to be provided in a “*consistent and clear*” format in a single location to reduce the work in gathering information. Participants spoke of requiring; “*less waffle, more facts and action plans*” and “*daily updates so we don’t have to research everything ourselves*”.

Participants perceived that they required support in the provision of information. Information that was required to maintain quality routine PHC care included clear patient management and infection control protocols and local information about “*updates as to contact tracing in our area*” and “*what is happening in local areas with regards to positive cases*”.

Greater communication through practical education sessions on COVID-19, screening procedures, infection control considerations and management of patients were seen as potential areas requiring support. Many participants also commented that the provision of education to the general public would support them to provide quality care. This included education to promote “*public awareness of risks*” and “*continued health promotion to the public not to become complacent*” about the spread of COVID-19. To ensure quality care was maintained, participants thought that a “*public campaign that it’s ok to see your nurse & doctor*” should be broadly communicated to “*let the community know we are still open and safe to come in*”. Finally, nurses were concerned about the impact of *“so called experts appearing on morning shows spouting their opinions”* and identified a need to reduce *“confusion in messages”*. Examples were provided of people being recommended to get their influenza vaccine but not supplying providers with sufficient stock to meet demand.

#### 3.2.3 Funding

One hundred and thirty four (11.0%) statements were concerned with the funding required to support the provision of quality PHC nursing care. The need for funded nurse-delivered telehealth was the most commonly reported statement regarding funding. The provision of government rebates for the delivery of telehealth services was seen crucial to both maintain patient care and enhance the retention of nurses. *“Billable items specific for nurses so they can provide Telehealth and optimal care while still bringing money into the practice*.*”*

Participants identified that in order to provide quality health care to the community, PHC nurses could do more in terms of consultations and assessments, home visits, chronic disease management and psychosocial issues related to COVID-19. However, funding for the provision of these services *‘billing for nurses’* was identified as an important support required to allow these services to be delivered. Some participants also mentioned the overall need for financial support by health services during COVID-19, including “*funds to keep us open”;* “*financial support for clinics that are struggling financially*”; and “*undertaking that current funding is not cut to the various primary health services*”. Other participants sought “*financial support to keep nurses employed at their regular hours or full time hours*” during the pandemic. This included both “*incentives for employers to maintain nurses*” and “*government relief packages for general practices to keep nurses in jobs*”.

#### 3.2.4 Industrial Issues

A number of issues were raised regarding threatened employment / lack of job security and employment conditions (fair pay and leave) that in general impact on the nurses ability to provide quality care during the pandemic. To provide quality care the nurses felt that they required *“guaranteed work hours are maintained”*, including a *“reinstatement”* or *“retention”* of work hours despite changes in the business models of their workplace due to social distancing requirements. If this was not possible, participants wanted to be able to access government income support schemes to cover lost wages. One participant summed up concerns regarding pay stating *“pay me properly and I won’t feel so used and abused”*.

Others added *“pay primary care nurses the same as hospital nurses” and “wages that reflect the skill, effort and risks undertaken by primary care nurses - currently my hourly rate is less than my 22yr old son’s. It’s demoralising”*. Some participants also identified differences in their ability to take paid leave in the PHC sector compared to the acute hospital. Such leave was sought if they might need to isolate themselves or care for family. Additionally, some participants felt that rewards or incentives in the form of *“hazard pay”*, a *“bonus”*, reductions in nurses’ registration fees or other professional fee reductions would support their practice.

#### 3.2.5 Self-care

A small number of statements (4%) recognised the psychological and emotional impact experienced by front line workers during a pandemic and the need for those nurses to engage in self-care strategies in order to stay well. Access to both informal and formal (funded) mental health services was perceived as a support required for the nurses to effectively engage in self-care to provide *“emotional support and to prevent burn out”* and thus remain well and functioning in the workplace. One participant stated that a key support is to *“ensure that the health and safety of health care workers is the number one priority so we can continue to care for the community”*. Achieving adequate self-care was seen to require *“down time with other colleagues”, “hours allocated to staff to maintain staff morale”* and a *“longer meal break time, time away from where I am working to debrief myself”*.

#### 3.2.6 Workplace factors

Forty five statements (3.7%) about supports required to maintain quality care related to factors associated with individual workplaces. The two key elements requiring support included improved local communication between team members and increased support from managers within the workplace. Communication areas included *“ongoing good information sharing by management”* and *“staff meetings to discuss as a group best plan moving forward”*. In terms of support from management, participants spoke of a need for *“real time support from organisation. Not support on paper. Doesn’t match reality”*. In contrast, nurses who experienced positive support and communication practices within their workplace such as *“regular meetings with area Exec* [*executive*]*”* described them as *“very helpful”*.

#### 3.2.7 Valuing nurses

Thirty eight statements (3.1%) focussed on how participants felt that greater acknowledgement of their value and contribution to the pandemic would support their practice. Participants sought recognition of the *“nurse role in education, reassurance and chronic disease management”, “the work that primary health nurses do during pandemics”* and *“that we actually do have skills and do our part”*. One participant stated *“we need to be valued to give care”*, while others observed that there is a need for *“GP’s to calm down and appreciate their nurses”* and for PHC nurses to *“not be treated in a condescending manner”*. Several participants felt that nurses were not included in the high level conversations around the health response to the pandemic, while others felt that nursing lacked leadership and “*representation at a national level to support primary health nurses in their roles”*.

## 4 DISCUSSION

This study was conducted at the height of the COVID-19 pandemic in Australia to understand the experiences of PHC nurses and highlight the supports required to facilitate the ongoing provision of quality nursing care. This is one of the few investigations specifically exploring the PHC nursing workforce during a pandemic (Halcomb et al., 2020). As such it provides an important insight into how community based nursing can be supported during such crises.

Supporting community based health services is essential to not only promote good infection control and case finding, but also to ensure that the community can continue to access healthcare for non-pandemic related health issues.

Our study highlighted a lack of personal protective equipment as the major concern for participants. In the context of COVID-19 a worldwide shortage of personal protective equipment, as a result of insufficient stockpiles and disrupted supply chains, saw health care professionals exposed to infection (Livingston, Desai, & Berkwits, 2020; Ranney, Griffeth, & Jha, 2020). This is not a new problem, with the supply of adequate personal protective equipment being reported during previous pandemics and epidemics (Cohen & Casken, 2011; Huang, Lin, Tang, Yu, & Zhou, 2020; Jones et al., 2017; Michaelis, Doerr, & Cinatl, 2009; Speroni, Seibert, & Mallinson, 2015). The significant number of health care workers dying globally due to COVID-19 exposure (Ehrlich, McKenney, & Elkbuli, 2020), highlights the need for more to be done internationally to ensure that sufficient personal protective equipment is available to protect health care professionals during future pandemics. In addition to addressing supply issues, clearly communicated and evidence-based principles around recommended personal protective equipment use will ensure prudent use of available supplies (Verbeek et al., 2020).

COVID-19 has tested international health care systems in a way not seen before in our lifetimes. This study highlighted that while participants experienced positive aspects around information sharing and communication, improvements in the delivery of messages, community and health professional education and clearer information about COVID-19 (including management protocols and up to date information) could support them to better deliver quality care. Similarly, in their systematic review of the pandemic experiences of acute care nurses, Fernandez et al. (2020) reported that nurses wanted to ensure that they had the appropriate education to provide care and felt that rapidly changing information increased their stress levels. Enhancing systems and processes for communication across the health system during crises is particularly important in PHC given the large numbers of individual health providers involved in the delivery of care.

The funding of nursing services in Australian PHC and industrial issues for PHC nurses are intertwined. In Australia, PHC nurses are employed in a range of small business general practices, non-government organisations and community care settings, as well as State funded community nursing services (Guzys et al., 2017). This creates complexity as the funding received for the delivery of nursing services is variable depending on the specific scheme which funds that service. Such variation has a flow on effect to create challenges in the consistency of remuneration and work conditions for nurses (Halcomb, Ashley, James, & Smyth, 2018). While the issue of funding nursing services in PHC was highlighted during the COVID-19 pandemic, it has been previously identified as a challenge to Australian PHC nurses providing quality care (Halcomb & Ashley, 2019). The issues raised by this study, and throughout COVID-19, highlight the urgent need to address funding and industrial concerns around PHC nursing to ensure that these nurses have job security and are retained in PHC employment (Australian College of Nursing, 2020; Halcomb et al., 2020).

This study identified concern for participants’ own mental health and well-being during and following the pandemic. The psychological impact of pandemics on acute care nurses has been previously identified (Chung, Wong, Suen, & Chung, 2005; Holroyd & McNaught, 2008; Shih et al., 2007). Many nurses have a strong sense of professional obligation to provide nursing care during a pandemic (Fernandez et al., 2020). However, the uncertainty of the situation, extreme pressures of high workload demands and need to prioritise resources, have been reported to create feelings of vulnerability, loneliness, powerlessness and both physical and psychological exhaustion (Fernandez et al., 2020; Usher, Bhullar, & Jackson, 2020). The impact of the nursing role is also overlaid with the psychological impacts facing all members of the community in terms of fear of contracting the pathogen, concerns for family and the impacts of social isolation (Shah et al., 2020; Usher et al., 2020). Ensuring that nurses, and other health professionals are well supported to maintain their mental health during times of crises is an important strategy to ensure that the workforce remains available to provide health care services to the community during and beyond the crisis.

Participants’ experiences of the COVID-19 pandemic raised the issue of Australian PHC nurses not feeling valued or recognised for their role. Perceptions of a lack of respect for nurses, and a lack of a voice and autonomy are cited as key factors leading to nurses leaving the profession (Sumner & Townsend-Rocchiccioli, 2003). Conversely recognising the efforts of nurses attracts nurses to work during future crises (Khalid, Khalid, & Qabajah, 2016). Despite significant advances in the professionalization of nursing, these study findings demonstrate that the male dominated medical profession is valued ahead of the female dominated nursing profession (Wynd, 2003). Communicating the value of PHC nursing in providing care is an ongoing issue in the sector, where more could be done to promote the role within the PHC team, the community and as contributors to health policy (Halcomb et al., 2018). This highlights the need for professional nursing organisations to take an active role to advocate on behalf of the profession and ensure that the voice of nurses is valued within the health care and health policy sphere across both acute and PHC.

## 5 LIMITATIONS

This study was conducted in rapid response to concerns raised by PHC nurses on social media as COVID-19 emerged in Australia. Given this unprecedented context, these data represent a snapshot of a single point in time and, as such, do not capture changes arising from the evolving pandemic response. Despite the significant survey response over a short timeframe, recruitment via social media may exclude nurses not present on these platforms. However the snowballing effect of recruitment cannot be ruled out as nurses on social media could have shared survey within their professional networks via email or word of mouth. In addition, lack of reliable data on the number of nurses employed in PHC settings precluded calculation of response rate. Similar sampling and recruitment limitations have been identified in previous research targeting this group (Halcomb, Salamonson, Davidson, Kaur, & Young, 2014). While findings revealed key supports required to retain and optimise the role of PHC nurses during a pandemic, further qualitative inquiry could have provided additional value.

## 6 CONCLUSION

This study provides new evidence about the support requirements of PHC nurses during a respiratory pandemic. As the threat of future pandemics looms internationally, these findings reveal areas in which support can ensure that PHC nurses can continue to provide quality care during the pandemic. Maintaining quality PHC nursing care has the potential to address many health and social issues within the community, thus reducing secondary morbidity and mortality and promoting health. As such the supports identified in this paper should inform pandemic planning into the future.

## 7 IMPLICATIONS FOR NURSING MANAGEMENT

PHC services are integral to the prevention, care and management of health care in the community, particularly during pandemics or other health crises. PHC nurses are frontline workers and require adequate support, but historically focus has been on acute care sector’s needs (Halcomb et al., 2020). Nursing management need to ensure that the needs of this workforce are addressed to ensure that the capacity to deliver high quality care is maintained in the community, to reduce secondary morbidity and mortality. COVID-19 has shown a responsive PHC nursing workforce where key issues about personal health and safety, care quality, and job security need to be addressed. However, nurse managers need to utilise strategies in the PHC context to support and manage risk for nurses and the community.

## Data Availability

Data is available by contacting the first author.

## Acknowledgements

We would like to thank the nurses who generously participated in the survey. Thanks also goes to the Australian College of Nursing and Australian Primary Health Care Nurses Association for supporting this work.

